# Projected geographic disparities in healthcare worker absenteeism from COVID-19 school closures and the economic feasibility of child care subsidies: a simulation study

**DOI:** 10.1101/2020.03.19.20039404

**Authors:** Elizabeth T Chin, Benjamin Q Huynh, Nathan C Lo, Trevor Hastie, Sanjay Basu

## Abstract

**Background:** School closures have been enacted as a measure of mitigation during the ongoing COVID-19 pandemic. It has been shown that school closures could cause absenteeism amongst healthcare workers with dependent children, but there remains a need for spatially granular analyses of the relationship between school closures and healthcare worker absenteeism to inform local community preparedness.

**Methods:** We provide national- and county-level simulations of school closures and unmet child care needs across the United States. We develop individual simulations using county-level demographic and occupational data, and model school closure effectiveness with age-structured compartmental models. We perform multivariate quasi-Poisson ecological regressions to find associations between unmet child care needs and COVID-19 vulnerability factors.

**Results:** At the national level, we estimate the projected rate of unmet child care needs for healthcare worker households to range from 7.5% to 8.6%, and the effectiveness of school closures to range from 3.2% (*R*_0_ = 4) to 7.2% (*R*_0_ = 2) reduction in fewer ICU beds at peak demand. At the county-level, we find substantial variations of projected unmet child care needs and school closure effects, ranging from 1.9% to 18.3% of healthcare worker households and 5.7% to 8.8% reduction in fewer ICU beds at peak demand (*R*_0_ = 2). We find significant positive associations between estimated levels of unmet child care needs and diabetes prevalence, county rurality, and race (*p* < 0.05). We estimate costs of absenteeism and child care and observe from our models that an estimated 71.1% to 98.8% of counties would find it less expensive to provide child care to all healthcare workers with children than to bear the costs of healthcare worker absenteeism during school closures.

**Conclusions:** School closures are projected to reduce peak ICU bed demand, but could disrupt healthcare systems through absenteeism, especially in counties that are already particularly vulnerable to COVID-19. Child care subsidies could help circumvent the ostensible tradeoff between school closures and healthcare worker absenteeism.

## Background

School closures are a common measure of pandemic mitigation for many countries, driven by the logic that social distancing reduces transmission^1–3^. Although school closures are known to reduce transmission, previous works have suggested that school closures could have downstream consequences on the healthcare system such as healthcare worker absenteeism^4,5^.

In the absence of a federal mandate for school closures, the decision of whether or not to close a school is determined by local authorities. However, lack of granular data has restrained previous studies to providing estimates based on state- or national-level data, underscoring the need for more detailed analysis^3^. The needs and capabilities of both schools and healthcare systems vary drastically across the United States, so county-level simulations of healthcare worker absenteeism and school closures could be more impactful and targeted for local communities than state- or national-level simulations. COVID-19 vulnerability factors such as social determinants of health (e.g. rurality, race) and complicating comorbidities, such as diabetes or cardiovascular disease^1^, vary geographically in the US, further highlighting the importance of regional analysis^7–9^.

To maintain healthcare systems in the event of a school closure, it could be beneficial to assist healthcare workers with child care. Previous work has shown that increased wages are associated with lower absenteeism, so it is possible that child care subsidies could reduce absenteeism by alleviating the financial burden of child care for healthcare workers as well as further incentivizing them to remain at work^10,11^. Furthermore, the costs of child care (which is the main barrier to finding child care) and the salaries of healthcare workers vary geographically, which would affect both the necessity and the economic feasibility of child care subsidization for healthcare workers in those areas^12^.

In previous works, Sadique et al and Lempel et al provided national level cost analyses of school closures under a variety of model assumptions and closure lengths^4,5^. Bayham and Fenichel provide state-level estimates and include a tradeoff analysis on whether closing schools reduces mortality after accounting for disruption to healthcare systems from absenteeism. Given the close tradeoff in mortality for school closures and absenteeism, it would be beneficial to explore ways to circumvent the ostensible tradeoff through child care subsidies^13^.

Here, we provide national- and county-level models that estimate rates of unmet child care needs for healthcare worker households in the event of school closure, the effectiveness of school closures by reduction of peak ICU bed demand, and the ecological association between COVID-19 vulnerability factors and estimated unmet child care needs. We also demonstrate the economic feasibility of child care subsidies as a measure to address healthcare worker absenteeism.

## Methods

### Data

To find county-level demographic and occupational data, we use 5-year estimates from the American Community Survey (ACS)^14^ and the Integrated Public Use Microdata Series (IPUMS)^15^, a database derived from ACS. The ACS provides comprehensive coverage of data at the county level across factors such as education, housing, employment, and income. For probability estimates of child care dependency, we use data from The National Household Education Survey and a Pew Research Center survey on working parents^12,16^. For county-level estimates of health assessments we use the Institute for Health Metrics and Evaluation and the CDC Diabetes Interactive Atlas^17,18^. For county-level fair market rent estimates, we use data from the U.S. Department of Housing and Urban Development (HUD)^19^. For child care cost estimates, we use data from Child Care Aware of America (CCAoA)^20^. We define healthcare workers as individuals belonging to the ACS categories of practitioners (e.g. physicians, nurses, technicians) or support staff (e.g. orderlies, aides, assistants).

### Population simulation

We simulate county-level demographic and occupational data for each county in the United States. We obtain estimates of the number of healthcare workers in each county and simulate distributions of them into gender and household type (no children, married with children, single male with children, single female with children) based on existing county-specific estimates from the ACS. We focus our analyses on households with children below age 13 - although children under the age of 5 do not attend school, daycare services would likely also be closed in the event of school closures.

We seed probabilities of being unable to find child care with data from NHES, Pew Research Center, the US Census Bureau, and IPUMS. Child care arrangements vary significantly based on parental employment, familial relations, between single and dual-parent households, and gender differences in caretaking of children^21^. In order to simulate which individual in a married couple would be responsible for child care in the event of a school closure, we draw upon survey data from both the Pew Research Center and the US Census Bureau indicating that 89% of working couples rely on the mother for primary child care^16^. We also test sensitivity by using an estimate of 60% instead of 89%.

To simulate ability to find child care in the event of a school closure, we test two different model assumptions:

1. Healthcare workers have difficulty finding child care at the same rates as national estimates. To simulate the probability a worker can find a child care alternative, we draw upon data from the NHES, which found that 50% of households had difficulty finding or could not find satisfactory child care.
2. Difficulty finding child care could be estimated from the household structure of healthcare workers. To simulate household statistics of healthcare workers, we use nationally representative microdata from IPUMS. We take employed healthcare workers who are either the head of the household or the partner of the head of the household and extract the age, relationship, and employment status of each member of the household. We estimate the ability to find child care by identifying other members of a household that could provide care. We define alternative child care as any member within the household that is over 13 and not employed (under 16, unemployed, or not in the labor force). We stratify the data by state, sex, occupation (practitioner or support staff), and partnership status (single or couple) to estimate the state-specific family structures of healthcare workers. We weight these state-specific derived rates of unmet child care need based on county-level demographic information to obtain estimates for each county.

Models under the first assumption may provide better estimates in that they include cases beyond household structure (e.g. child care from a relative living elsewhere), but are limited by the assumption that healthcare workers have the same difficulties finding child care as the national average. Models under the second assumption may provide better estimates in that they account for child care difficulties specific to healthcare workers, but are limited by the assumption that all possible caregivers live in the same household as the child.

### Estimating unmet child care needs

We estimate the rate of unmet child care needs for healthcare workers over each county in the United States. Using the probabilities determined in the previous step, we simulate whether or not a given health care worker will be able to find alternative child care in the event of a school closure. At both the national- and county-level, we draw 1000 simulations from multinomial distributions. We determine unmet child care needs by simulating whether a healthcare worker is the primary caregiver of a household, and whether they are able to find alternative child care in the event of a school closure. We calculate rates of unmet child care needs by dividing over the total number of healthcare workers.

We then repeat the above steps across healthcare worker subgroups (practitioner or support staff) to get a range of estimates. We also perform different estimations based on the different model assumptions proposed in the previous section.

### Transmission models

We model the impact of school closures by county using a compartmental model with an age-structured SEIR framework^22^. We divide the population into four age groups: 0-19 years, 29-39 years, 40-59 years, and 60+ years. Transmission events occur through contact between susceptible and infectious individuals. Since rates of contact differ between age groups, we construct a WAIFW (Who Acquires Infection From Whom) matrix from non-physical and physical contact data between age groups^23^. We assume that social distancing will result in a 50% reduction of interactions and school closures will result in a 90% reduction in interactions among children^24^. Since increased household interactions is often cited as an unintended side effect of school closures^25,26^, we also increase interactions between children and other age groups by 10%.

We assume an incubation period of 5.1 days and an infectious period of 6.5 days^26,27^. The *R*_0_ of SARS-CoV-2 is estimated to be between 2.0 and 6.0, and we examine values within that range (*R*_0_=2.0, 2.5, 3.0, 3.5, 4.0, 4.5, 5.0, 5.5, 6.0). Since the COVID-19 outbreak curve is over a short duration, we ignore births, death, and immigration. We assume that 86% of infections are mild or asymptomatic, with asymptomatic individuals as 50% less infectious than symptomatic individuals^28^. Symptomatic individuals are assumed to reduce contact by 75%. Age-stratified hospitalization rates and infection fatality ratios were obtained from Verity et al^29^. We choose to only apply the infection fatality ratios to symptomatic individuals to obtain a conservative estimate. We assume that individuals develop immunity after recovering from COVID-19 in the short term. To estimate the demand on the healthcare system, we assume that 30% of hospitalizations will require critical care (invasive mechanical ventilation, vasopressor support, or further intensive care-level intervention), and that individuals requiring hospitalization will stay for 8 days and individuals requiring critical care will stay in the ICU for 10.4 days^26^. To simulate the effects on a particular county, we seed the simulation for county age demographics. We estimate the effectiveness of school closures by calculating the reduction in peak ICU bed demand between social distancing and social distancing plus school closure conditions.

### Regression analysis

We perform multiple ecological regression analysis to find associations between unmet child care needs and COVID-19 vulnerability factors. We use a quasi-Poisson regression model with rates of unmet child care needs as the outcome, and healthcare worker population as weights^30^. Our factors of interest, based on available county data, are diabetes prevalence, cardiovascular disease mortality, and rurality. We control for race, age, state, household status, sex, population, and fair market rent. We run separate models for cardiovascular disease, diabetes, and rurality, as well as one for controls only.

### Economic analysis

We calculate the economic costs of healthcare worker absenteeism from school closures and compare them to the costs of providing child care to healthcare workers with children. We estimate the percentage of absenteeism from school closures as the percentage of unmet child care needs multiplied by a constant *ρ*, since there may be overlap in absenteeism from other factors (e.g. sick leave) or attendance despite unmet child care needs. We vary *ρ* = {0.5, 0.6, 0.7, 0.8, 0.9, 1} to get a range of estimates. We estimate the cost of absenteeism as worker wages multiplied by number of workers (split by gender and practitioner/support staff subgroups) within a county, multiplied by 1.4 to account for value not captured by wages, such as taxes, pension, cost of overtime, paid sick leave, etc^24^.

We estimate the cost of providing child care to healthcare workers with children by estimating county-level child care costs and the number of healthcare workers with children per county. We estimate county-level child care costs for both full-time and part-time child care with a method similar to that used by the Economic Policy Institute’s Family Budget Calculator (see Supplement)^31^.

To compare the two costs, we divide the cost of healthcare worker absenteeism from school closure by the cost of providing child care to all healthcare workers with children at the county level to get a coefficient *ω*. We then calculate the percentage of counties with *ω* > 1 at each level of *ρ*, indicating the percentage of counties where it is cheaper to provide child care to all healthcare workers with children than it is to bear the costs of healthcare worker absenteeism from school closure.

## Results

### Estimating unmet child care needs

Our national level simulation based on NHES data provided unmet child care needs estimates of 7.5%, 7.2%, and 7.9% for all healthcare workers, healthcare practitioners/technicians, and healthcare support staff, respectively. Our simulation based on IPUMS data provided higher estimates of 8.6%, 9.2%, and 7.4%. Our county-level approach revealed substantial variation in estimated healthcare worker unmet child care needs across counties, ranging from 2.0% to 18.6% (Figure 1).

**Figure 1.**
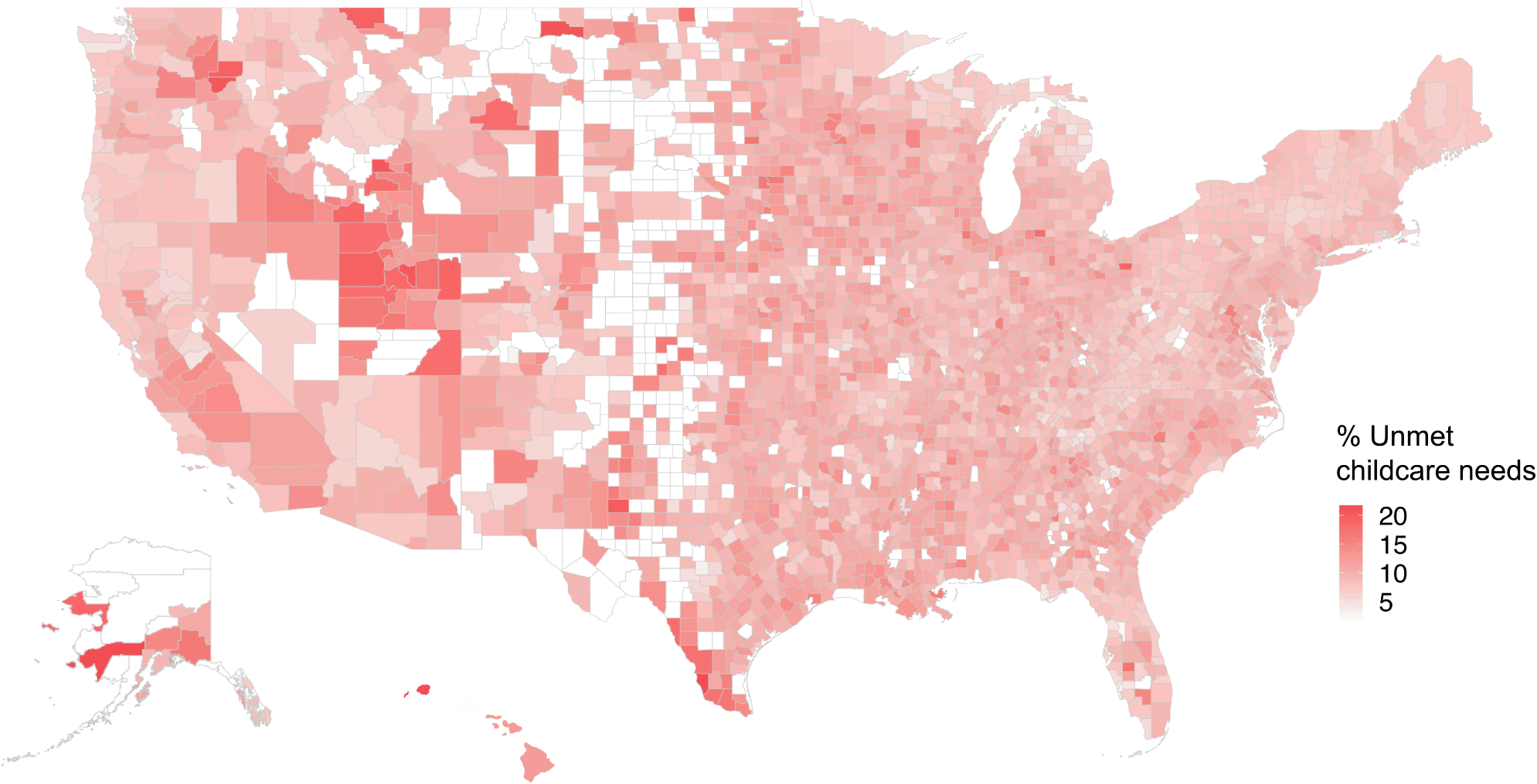
Estimated rates of unmet child care needs for healthcare worker households by county. Counties with confidence interval sizes in the 90th percentile or below (< ±5.95%) are shown.

### Transmission models

When assuming *R*_0_ = 2.0, our national-level SEIR model estimated a 7.38% and 7.22% reduction in peak demand for hospital beds and ICU beds, respectively. Our county-level estimates showed a reduction in peak hospitalization and ICU rates for all counties under school closure conditions, with substantial variation in hospital demand across counties, ranging from a reduction in peak demand of 5.87% and 9.08% for hospital beds and 5.75% and 8.83% for ICU beds (Figure 2). Our sensitivity analyses show the effectiveness of school closures decreases with increasing *R*_0_ values, which is consistent with past findings^32^. We observe from our models a reduction in hospital demand as a result of school closures with and without increased household interactions (Supplement Table 2).

**Figure 2.**
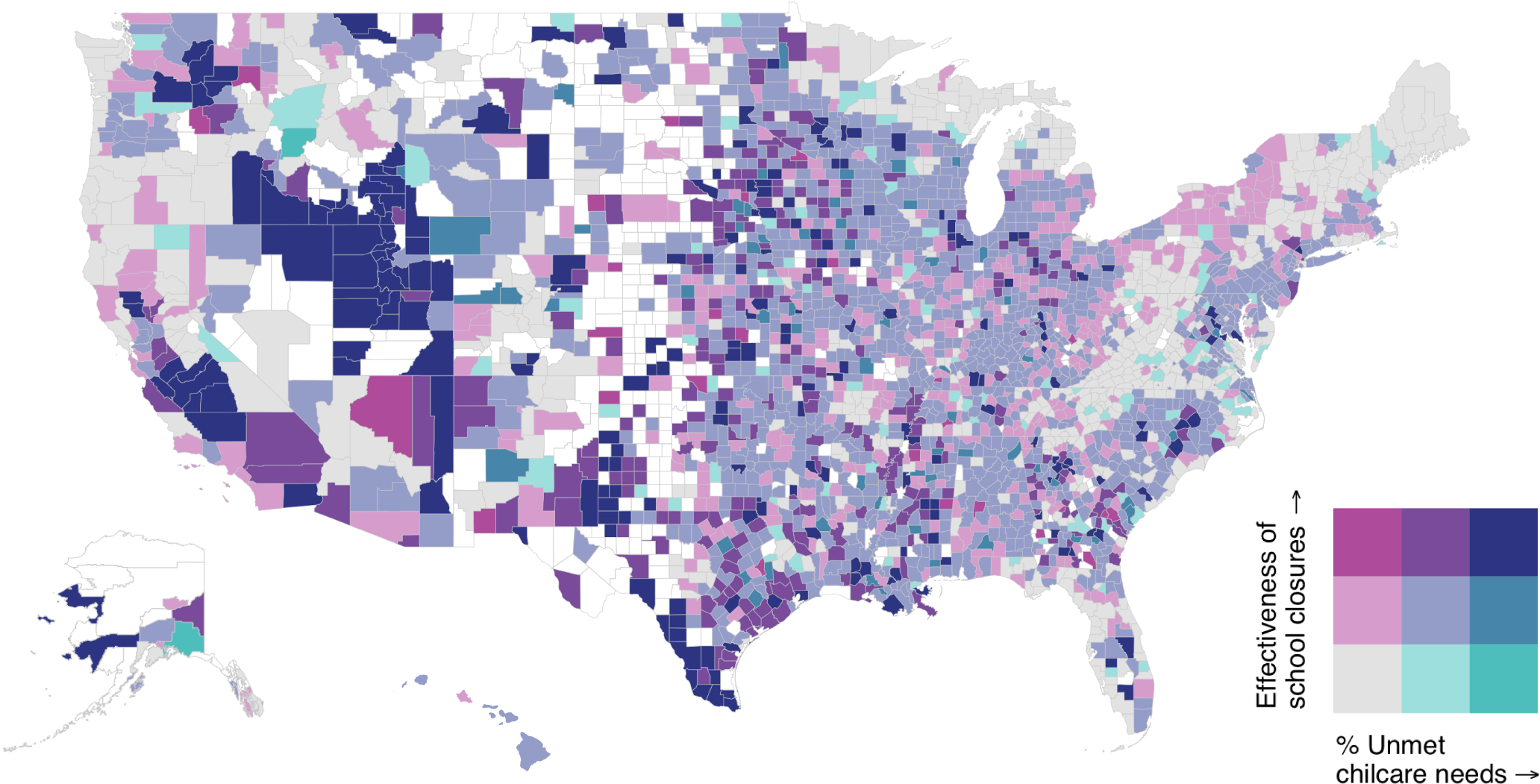
County-level comparison of percent of healthcare worker households with unmet child care needs and effectiveness of school closures using estimated reduction of peak ICU bed demand. Counties with confidence interval sizes in the 90th percentile or below (< ±5.95%) are shown.

### Regression analysis

We found from our regression analysis that diabetes prevalence is positively associated with unmet child care needs with a coefficient of 0.22, meaning a 1% increase in diabetes prevalence is associated with a 0.22 percentage point increase in healthcare worker households with unmet child care needs. Cardiovascular disease mortality is negatively associated with unmet child care needs, with a coefficient of less than *−* 1.859 *×* 10^4^. Rurality proportion has a positive coefficient of 0.02, so an increase from nonrurality to full rurality is associated with a 2 percentage point increase in healthcare worker households with unmet child care needs. Proportion of African Americans and proportion of Hispanics have positive coefficients of 0.04 and 0.15, respectively. (Supplement Table 3)

### Economic analysis

Based on our values of *ρ*, we estimated that for 71.1% to 98.8% of counties, it would be less expensive to provide child care subsidies to all healthcare workers with children than to bear the costs of healthcare worker absenteeism during school closures (*ω* > 1). The rurality proportions across counties remains relatively constant across values of *ρ*. (Figure 3)

**Figure 3.**
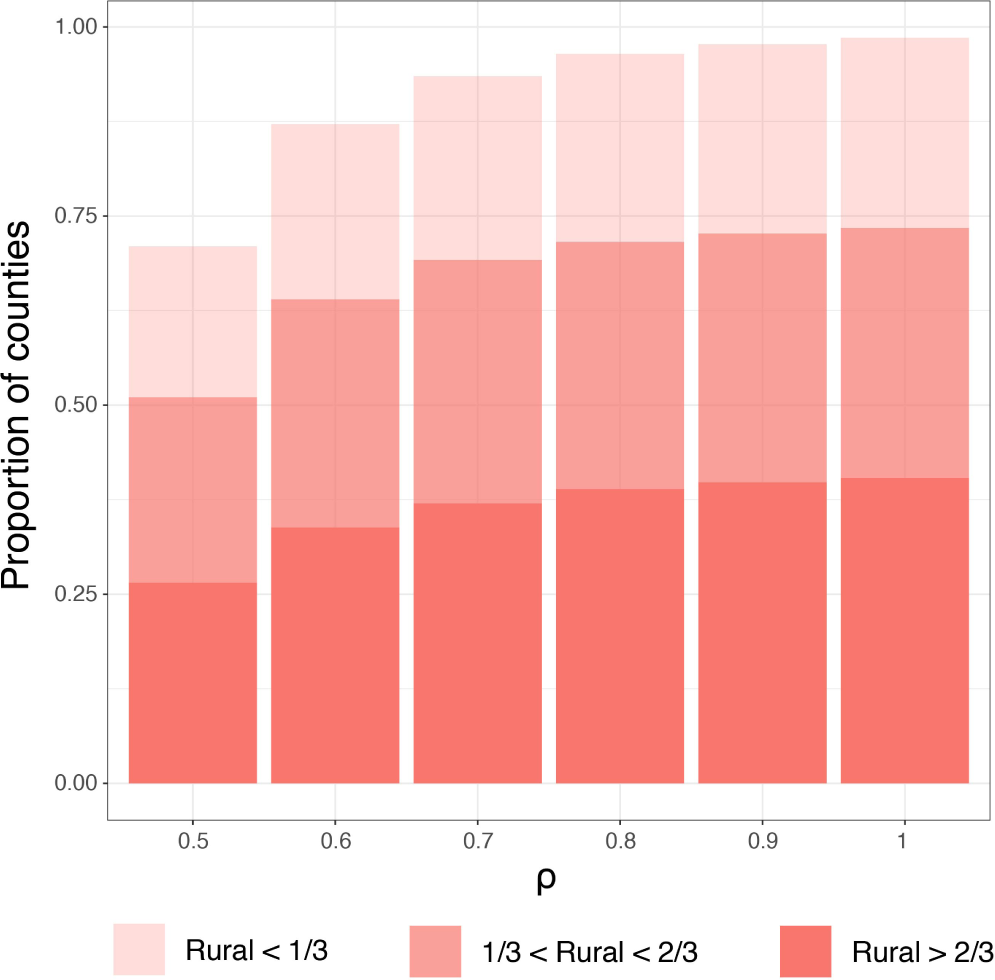
Proportion of counties with higher rates of lost wages due to absenteeism than costs of child care (*ω* > 1) across *ρ* = {0.5, 0.6, 0.7, 0.8, 0.9, 1.}. Bars are shaded based on the level of rurality of counties.

**Figure 4.**
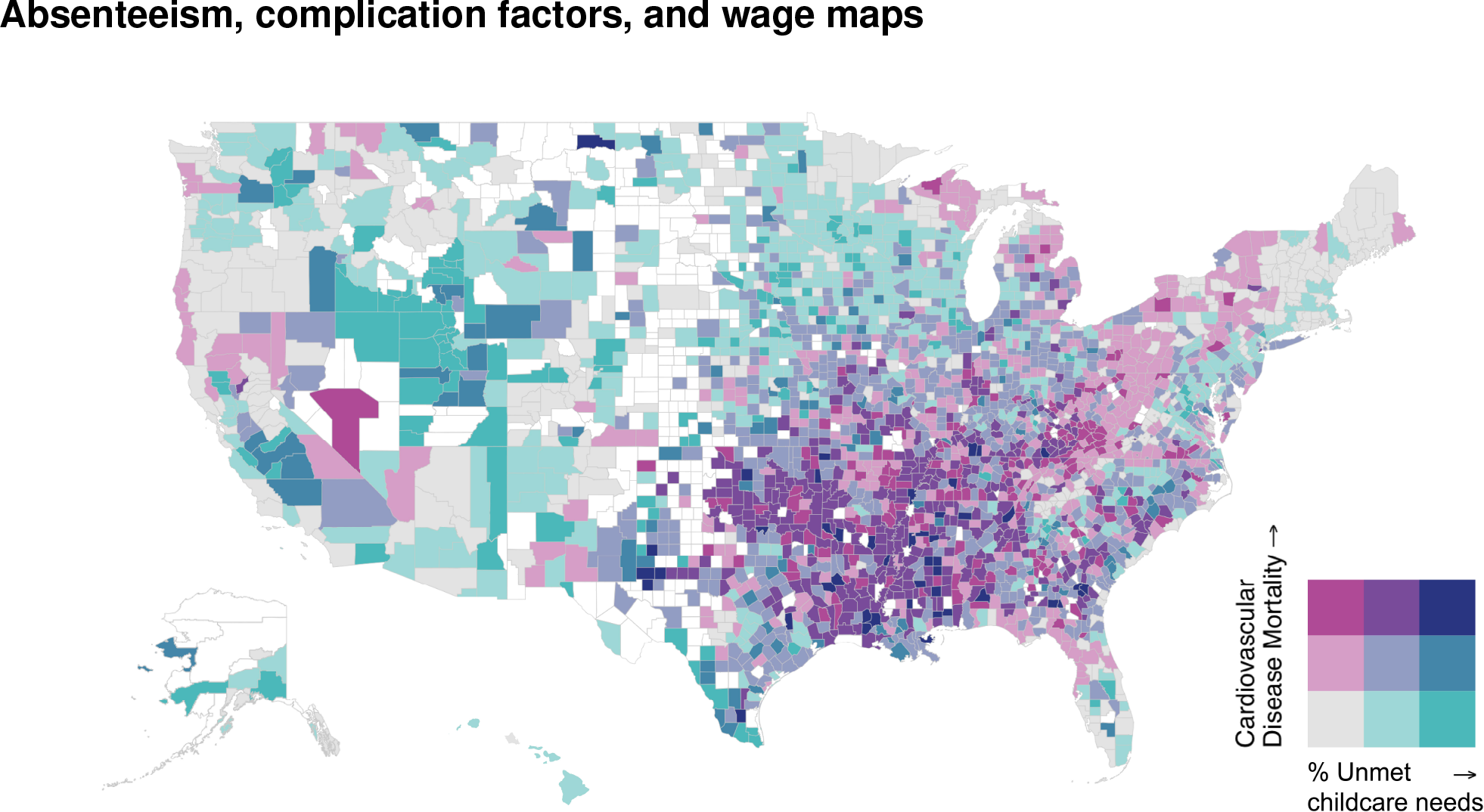
County-level comparison of percent absenteeism and cardiovascular disease mortality (deaths per 100,000 people). Counties with confidence interval sizes in the 90th percentile or below (< ±5.95%) are shown.

**Figure 5.**
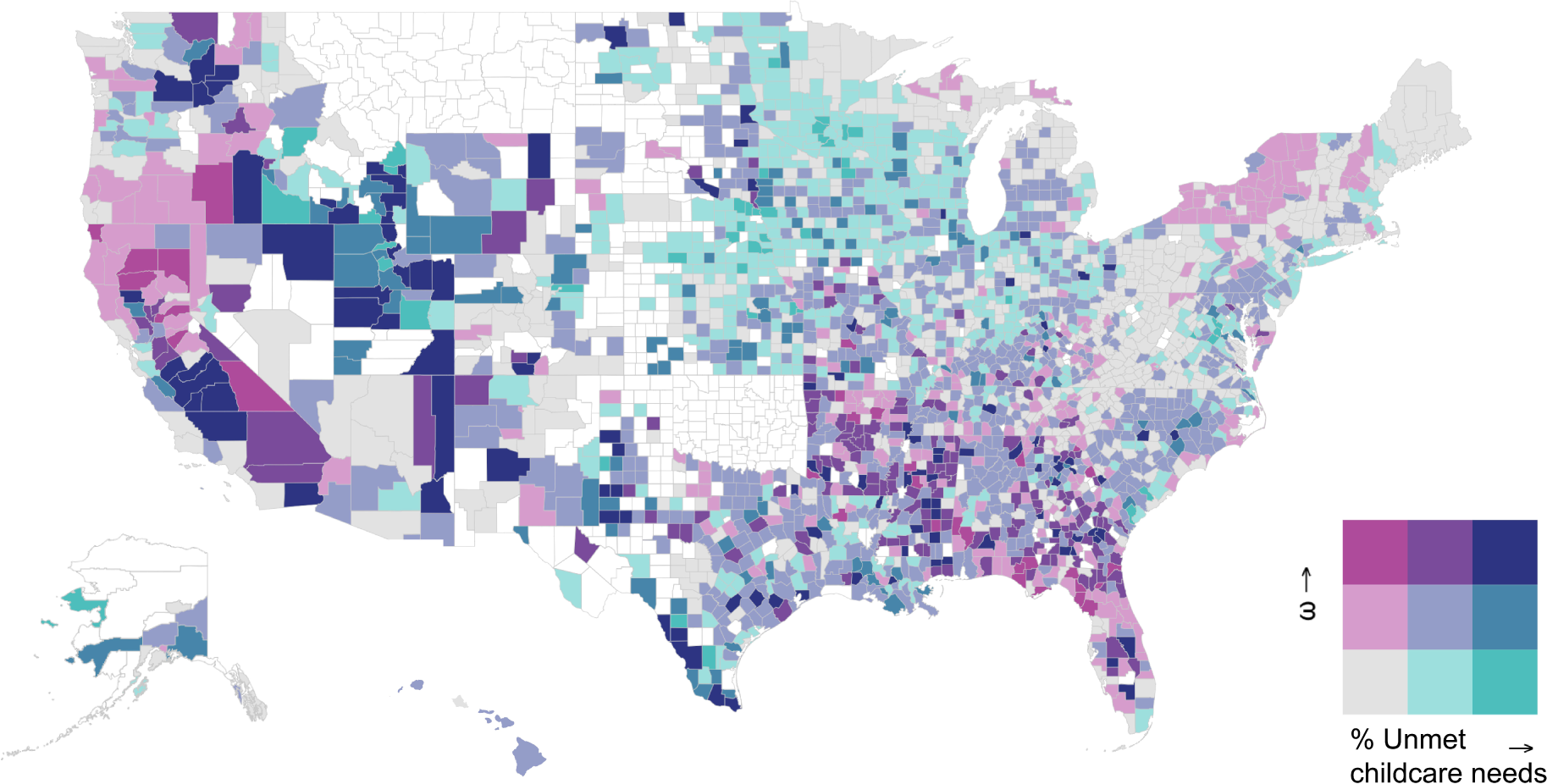
County-level comparison of percent absenteeism and *ω*. Counties with confidence interval sizes in the 90th percentile or below (< ±5.95%) are shown.

SEIR Equations

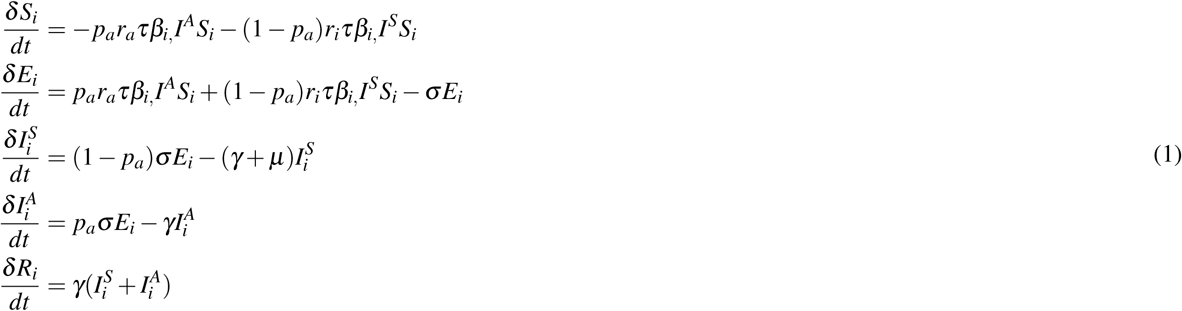

**Figure 6.** Differential equations for SEIR models. *i* is the age group, *p*_*a*_ = 0.83 is the proportion of infected that are asymptomatic. *r*_*a*_ = 0.5 is the reduction of infectiveness of an asymptomatic individual. *r*_*i*_ = 0.25 is the reduction in interaction of a symptomatic individual. *β*_*i*_ is age stratified contact rates derived from a WAIFW matrix, *τ* is the probability of transmission given contact derived from *R*_0_. The average length of incubation was set to 1*/s* = 5.1 days and the average length of infections was set to 1*/γ* = 6.5 days.

### Aggregate analysis

We observed a number of counties that could be viable targets for child care subsidies based on our estimates (Table 1, Figure 3). Counties like Conecuh County, Alabama and Todd County, South Dakota, have high rates of diabetes, rurality, projected unmet child care needs, as well as a high *ω*, suggesting that they would suffer disproportionately from COVID-19 in the event of school closures, but also that a child care subsidy would be relatively inexpensive for them. Similarly, Hidalgo County, Texas and Fresno County, California have high projected rates of unmet child care needs and *ω*, suggesting they are viable targets for child care subsidies. San Francisco County, California is one of the few counties with *ω* < 1 (due to high child care costs, low wages, and low projected unmet child care needs), illustrating the variance of our estimates within states. Counties like Bronx County, New York that have high projected school closure effectiveness but also high projected unmet child care needs, could also consider child care subsidies given the large estimated benefit of school closures.

**Table 1.** Estimated proportion of healthcare worker households with unmet child care needs, *ω*, closure effectiveness (CE), and actual diabetes prevalence for example counties. Closure effectiveness is defined as the percent reduction in peak ICU bed demand.

| State | County | Unmet childcare needs | $\omega$ | CE | Diabetes | Rural |
| --- | --- | --- | --- | --- | --- | --- |
| AL | Conecuh | 0.13 (0.09, 0.17) | 4.13 | 7.31% | 0.20 | 0.81 |
| CA | Fresno | 0.13 (0.12, 0.13) | 5.76 | 7.69% | 0.10 | 0.11 |
| CA | San Francisco | 0.04 (0.03, 0.04) | 0.78 | 6.39% | 0.08 | 0.00 |
| NY | Bronx | 0.11 (0.11, 0.12) | 1.33 | 7.63% | 0.13 | 0.00 |
| SD | Todd | 0.19 (0.11, 0.26) | 5.83 | 8.79% | 0.15 | 1.00 |
| TX | Hidalgo | 0.17 (0.17, 0.17) | 4.62 | 8.28% | 0.10 | 0.05 |

**Table 2.** Sensitivity analysis of absenteeism estimate using various population seeds. National Household Education Surveys Program (NHES) found that 50% of households had difficulty finding or could not find satisfactory child care. Integrated Public Use Microdata Series (IPUMS) are state specific seeds derived from the household structure of healthcare workers. Survey data from both the Pew Research Center and the US Census Bureau indicating that 89% of working couples rely on the mother for primary child care. To test sensitivity, we calculated absenteeism by assuming that 60% of working couples rely on the mother for primary child care.

Absenteeism estimate
| Population seeds | All | Practitioners/Technicians | Support staff |
| --- | --- | --- | --- |
| $NHES_{0.89}$ | 7.5% | 7.2% | 7.9% |
| $IPUMS_{0.89}$ | 8.6% | 9.2% | 7.4% |
| $IPUMS_{0.6}$ | 7.3% | 8.3% | 6.3% |

**Table 3.** Sensitivity analysis of transmission models under varying *R*_0_ values and contact conditions. School closures (SC) reduce the risk of child-child interactions by 90%. Household (HH) interactions increase child-other age group interactions by 10%. Both models assume social distancing, which reduces all interactions by 50%

| $R_0$ | Hospital beds: SC + HH | Hospital beds: SC | ICU beds: SC + HH | ICU beds: SC |
| --- | --- | --- | --- | --- |
| 2.0 | 7.38% | 8.19% | 7.22% | 8.01% |
| 2.5 | 5.91% | 6.6% | 5.87% | 6.32% |
| 3.0 | 4.71% | 5.26% | 4.58% | 4.94% |
| 3.5 | 4.16% | 4.5% | 3.98% | 4.3% |
| 4.0 | 3.49% | 3.88% | 3.25% | 3.56% |
| 4.5 | 3.23% | 3.5% | 3.01% | 3.23% |
| 5.0 | 2.79% | 3.19% | 2.54% | 2.87% |
| 5.5 | 2.43% | 2.75% | 2.29% | 2.47% |
| 6.0 | 2.31% | 2.44% | 2.16% | 2.29% |

**Table 4.**
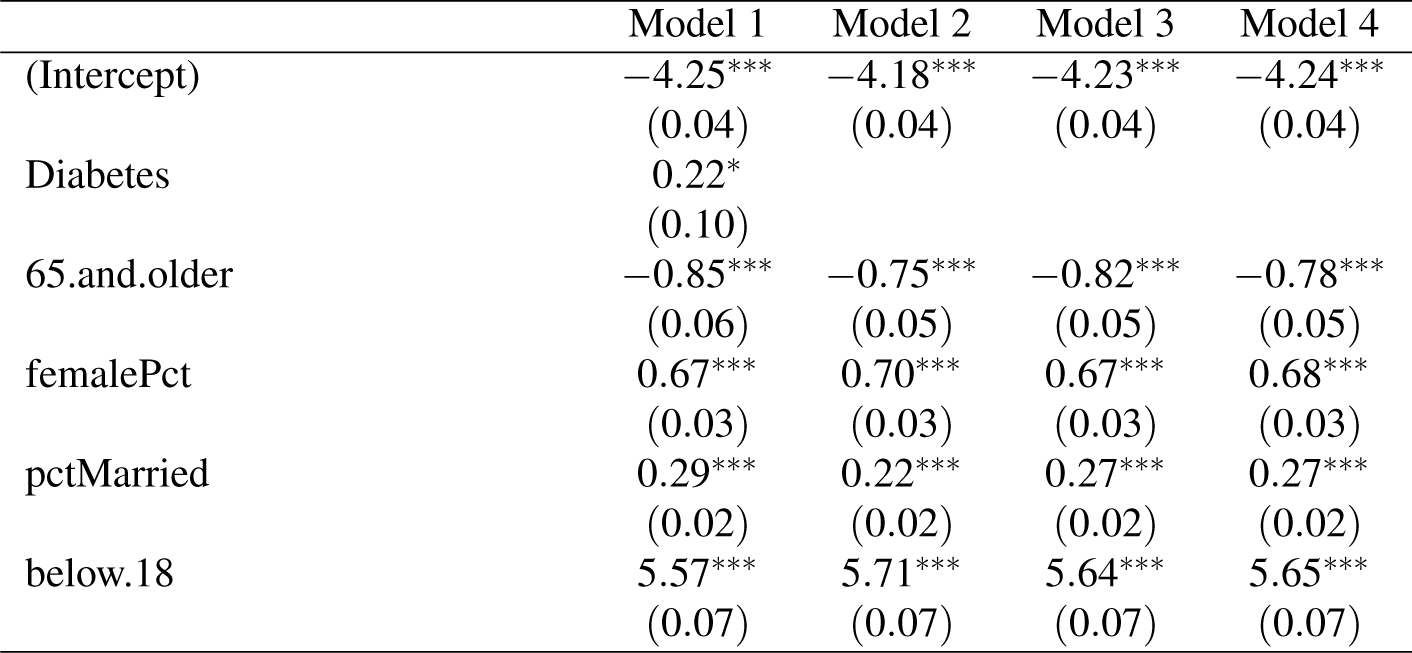
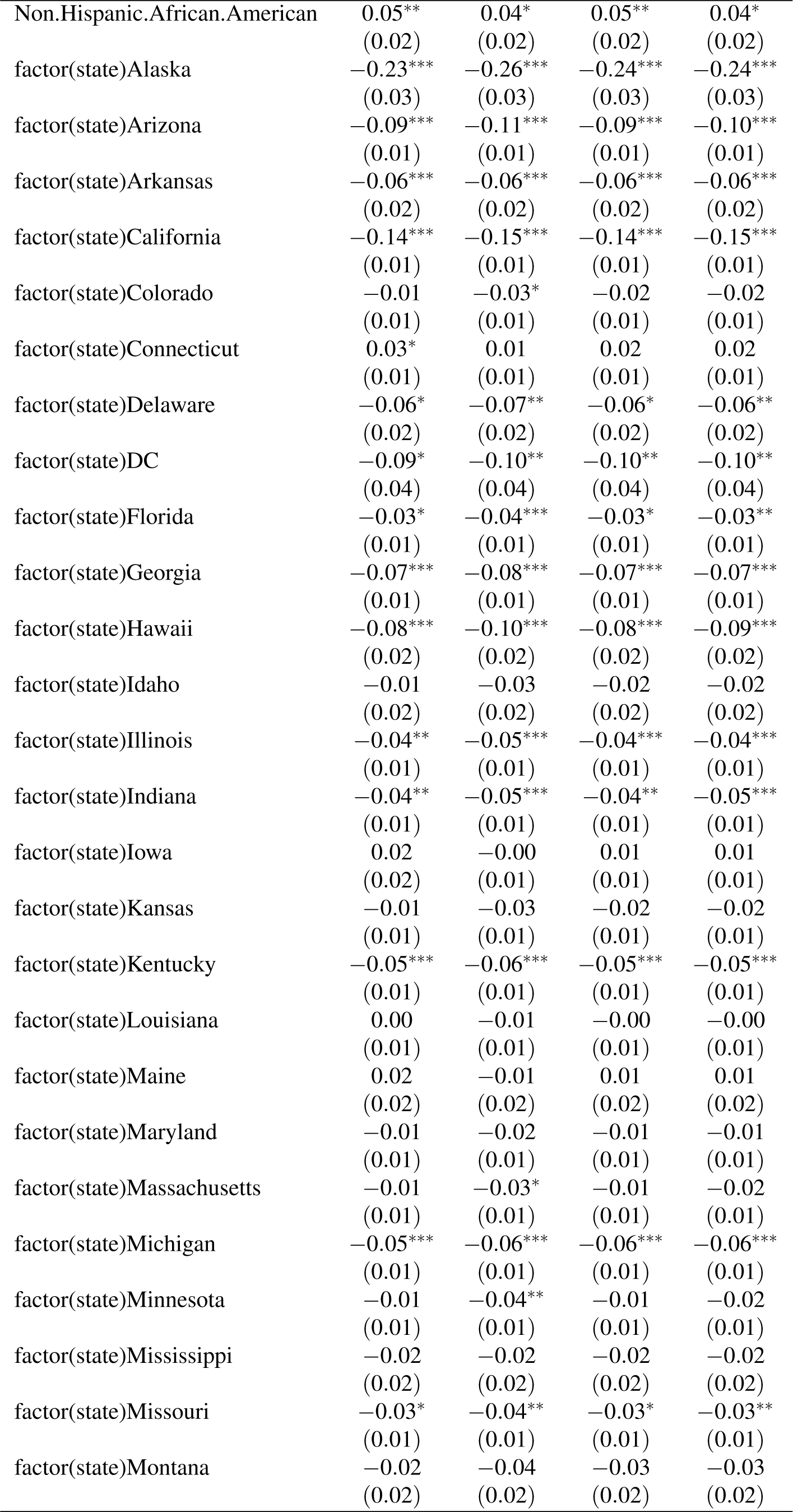
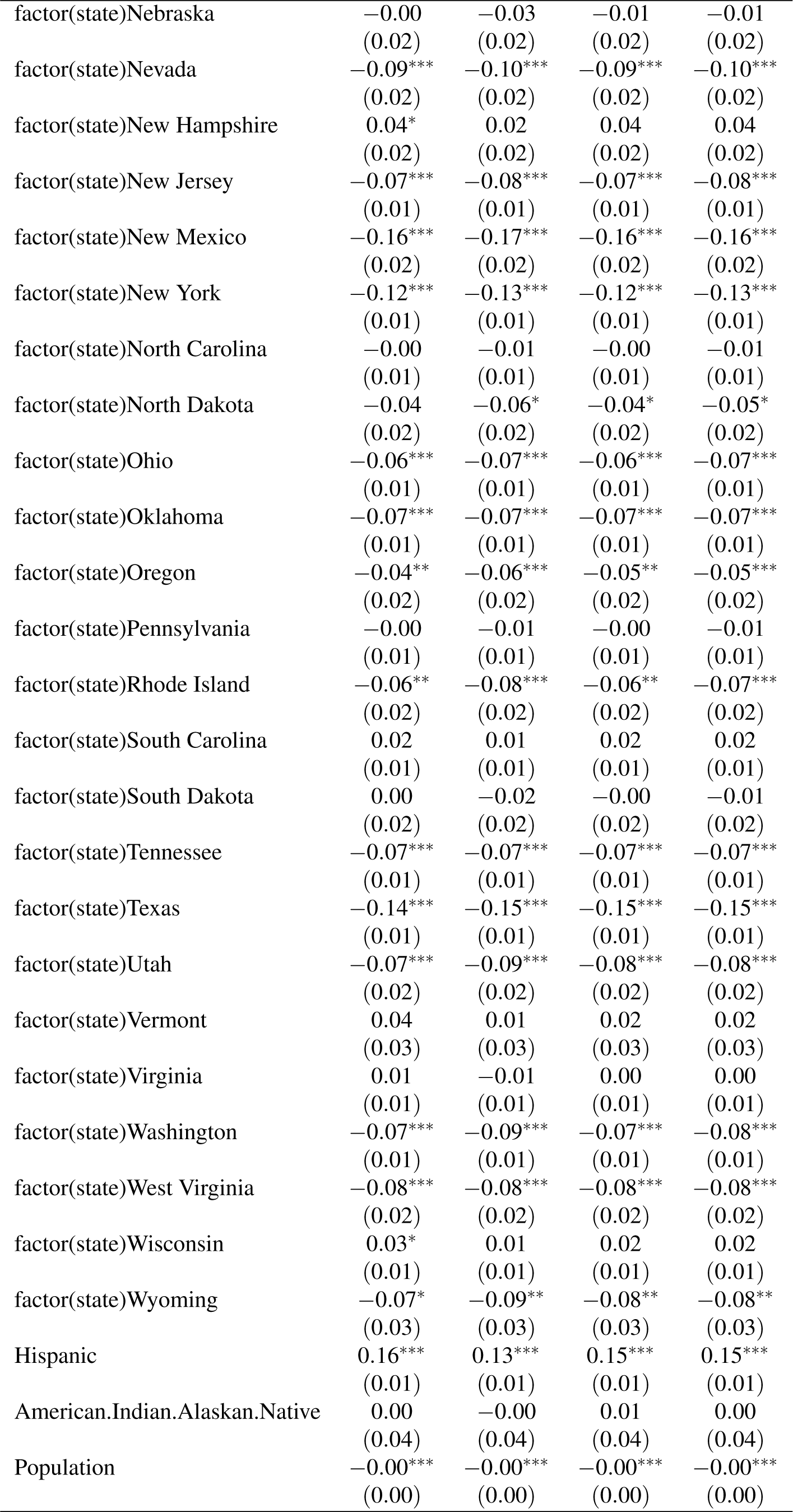
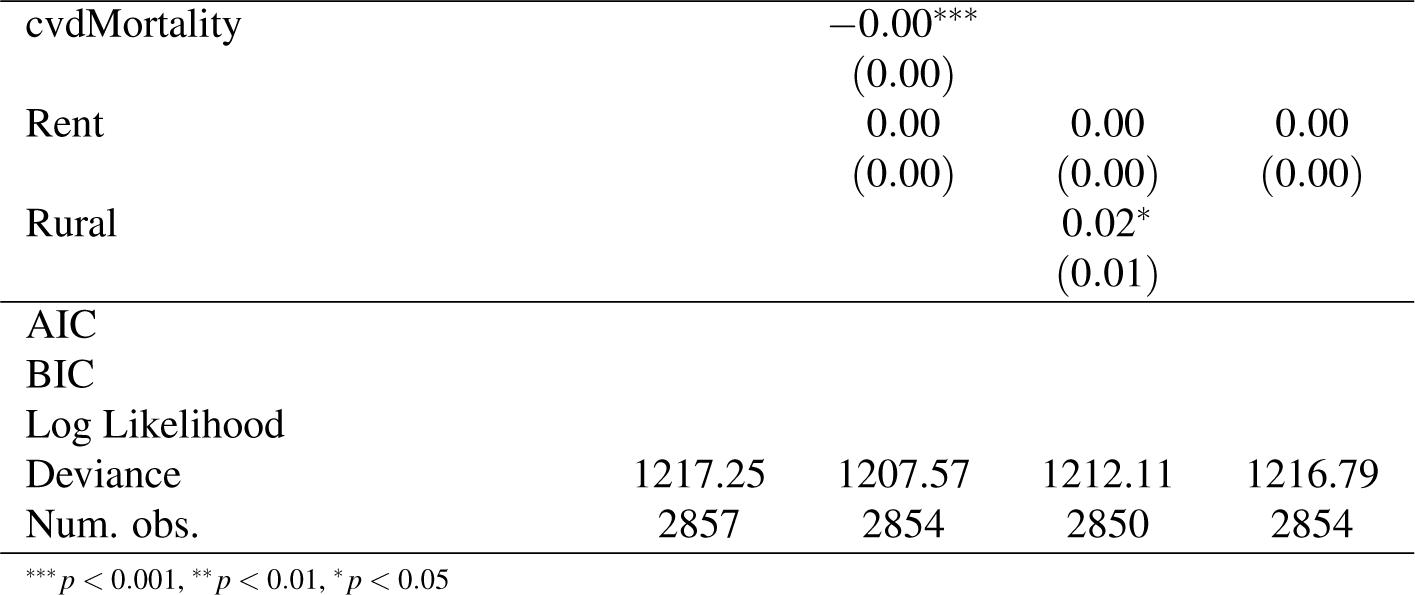
Regression output for models on diabetes, cardiovascular disease, percent rural, and controls.

## Discussion

Our models estimated generally high rates of unmet child care needs across different assumptions (> 7%), and our transmission models projected reduced peak ICU bed demand from school closures. However, since it is highly likely that hospitalizations and ICU bed demand would still far exceed bed capacity for many hospitals^26^ despite the effectiveness of school closures, we observe a need for an intervention to reduce absenteeism in the event of school closures. Because we observed large variance of our estimates between counties for all of our county-level analyses, identifying needs and interventions at the county-level is likely to be more effective at mitigating harm than a nation- or state-wide strategy.

Our regression analysis estimated that counties with higher percentages of diabetes prevalence, rurality, and Black/Hispanic populations would also have higher rates of unmet child care needs from school closures. Early data have shown that patients with diabetes have higher COVID-19 mortality rates^6^, and that African Americans are disproportionately represented in COVID-19 death counts.^2^ Furthermore, rural counties are more likely to lack adequate hospital capacity than urban counties.^33^ Without a way to mitigate absenteeism, counties that are likely to be most vulnerable to COVID-19 are also estimated to be more vulnerable to absenteeism from school closures, illustrating exacerbated geographic disparities in the absence of adequate child care.

To identify a potential approach to reducing absenteeism, we estimated that a majority of counties (71.1% to 98.8%) could save money by providing child care to their healthcare workers with children in the event of a school closure (*ω* > 1). Although it is likely that many child care avenues would also be closed in the event of school closures, subsidized child care costs could still prevent absenteeism by (1) incentivizing work attendance with extra wages, and (2) alleviating the financial burden on the entire household, enabling other family or household members to participate in child care.

As a simulation study, there are important limitations to our analysis. Simulations rely on assumptions to make predictions, and ours use assumptions derived from available data. For example, we do not know the number of healthcare workers with dependents - we estimate this based on representative data that could be inaccurate for some regions. Similarly, there are no datasets that tell us how many healthcare workers would be unable to find child care in the event of school closures - we instead estimate this using representative microdata. Lack of available data prohibits us from making precise estimates for counties with small populations. Given the current uncertainty of transmission parameters, our transmission models should not be used to accurately predict infection and hospitalization rates, but rather to estimate the relative effectiveness of school closures based on the age-demographics of each county. Although our economic analysis demonstrates the affordability of a child care subsidy, our method does not prove that child care subsidies would necessarily reduce absenteeism resulting from school closures. We emphasize that our work does not argue for or against school closures due to currently unclear fatality and transmission data, but rather that we highlight areas that would suffer more in the event of school closure and could therefore benefit more from child care subsidies.

Further research should investigate whether child care subsidies for healthcare workers would reduce absenteeism in the event of school closures from a pandemic. Additionally, research efforts should identify how school closures in pandemics impact more vulnerable populations for whom robust data does not currently exist. Further research efforts should also be placed to determine the effect of school closures on the absenteeism of other kinds of essential workers, instead of just healthcare workers.

## Conclusion

Our analyses suggest geographic disparities in unmet childcare needs of healthcare workers from school closures, exploring the possibility of targeted child care subsidies for local communities. We demonstrate the economic feasibility of child care subsidies to circumvent the tradeoff between school closures and healthcare worker absenteeism in the majority of US counties. Child care subsidies may play a critical role in maintaining the healthcare work force, and such actions could help reduce preventable harm resulting from school closures. Our study provides a step towards informing future work on better understanding the effects of locally targeted nonpharmaceutical interventions in the event of disease outbreak.

## Data Availability

All code and data will be made publicly available.

## Acknowledgments

We thank Dr. Euan Ashley (Stanford University) for his helpful comments on our manuscript. This material is based upon work supported by the National Science Foundation Graduate Research Fellowship under Grant No. DGE – 1656518 and the National Library of Medicine under Training Grant T15 LM 007033.

## Author contributions

ETC and BQH conceived and designed the study, conducted the analysis, and drafted the manuscript. NCL provided clinical perspective. TH assisted with the statistical analysis. SB supervised the study. All authors discussed and contributed to the final version of the manuscript.

## Supplement

### Economic analysis

Here we describe how we obtained county-level estimates of childcare costs and wages. We use state-level child care costs from CCAoA and adjust them to county-level by applying the ratio between state-level and county-level fair market rents from HUD. We calculate state-level rents from HUD by taking population-weighted averages of county rents. To estimate the number of healthcare workers with children at the county-level, we take the state-level proportion of healthcare workers with children from IPUMS and apply it to the county-level number of healthcare workers from ACS. We then calculate the county-level cost of providing child care to healthcare workers by multiplying child care costs by the proportion of healthcare workers with children.

For estimating county-level wages, some counties with low populations had redacted wages to preserve anonymity. We used multiple imputation by chained equations to impute these cases. To get all county-level wages, we multiplied the number of healthcare workers (by occupation group and sex) by their subgroup-respective county-level median wages.

## Sensitivity analyses

### Absenteeism estimate

### Transmission models

## Model output

The glm() calls for our models and model output are below.

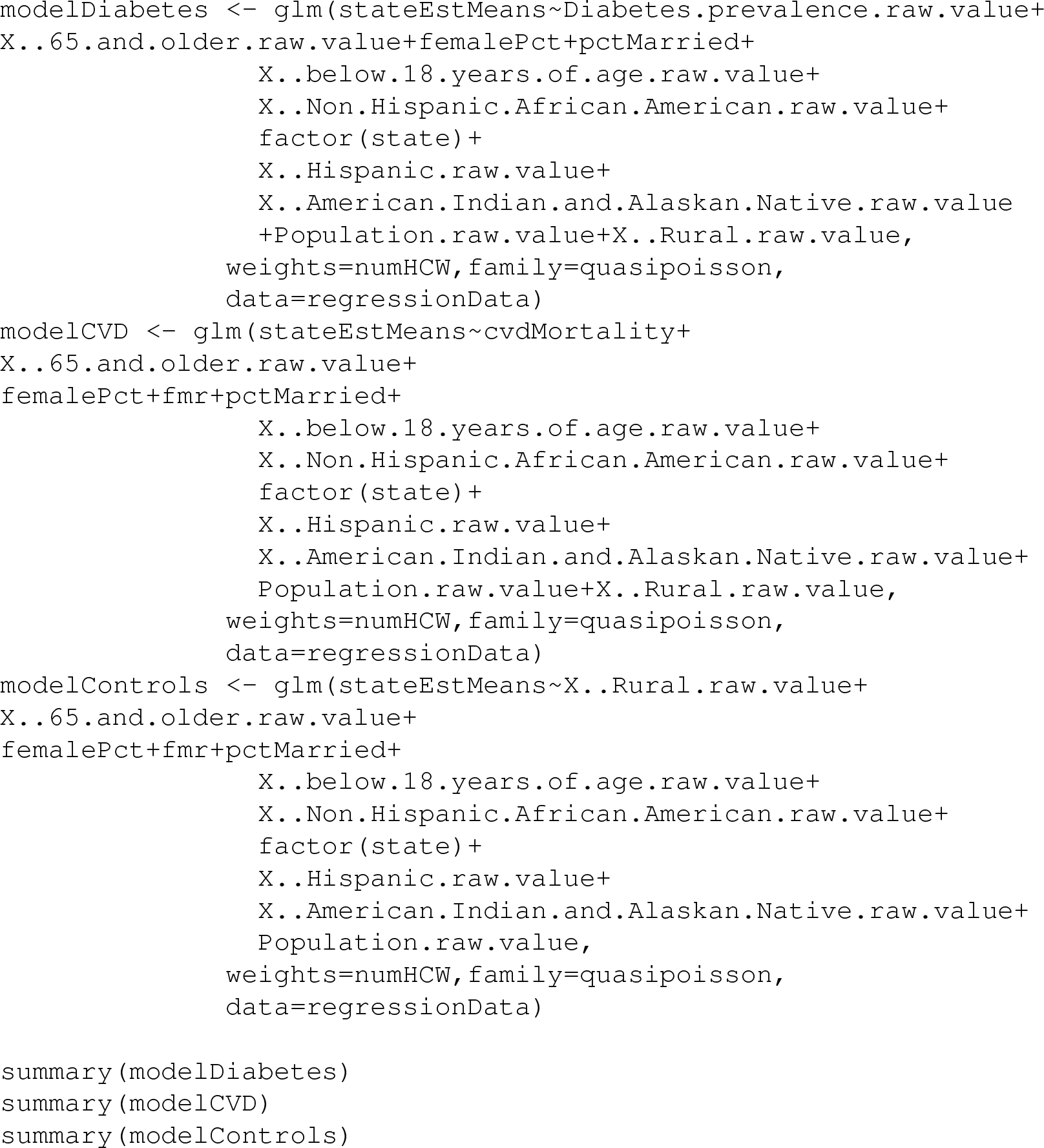

## Footnotes

1 China’s Center for Disease Control reported a 0.9% case fatality with no reported comorbidities and 7.3% and 10.5% for comorbidities of diabetes and cardiovascular disease, respectively.^6^

2 COVID-19 Dashboard. Milwaukee County, Wisconsin. https://county.milwaukee.gov/EN/COVID-1934. COVID-19 Statistics. Illinois Department of Public Health.http://www.dph.illinois.gov/covid19/covid19-statistics35. COVID-19 North Carolina Dashboard. North Carolina Department of Health and Human Services.https://www.ncdhhs.gov/divisions/public-health/covid19/covid-19-nc-case-count

